# Heterologous SARS-CoV-2 IgA neutralising antibody responses in convalescent plasma

**DOI:** 10.1101/2022.02.06.22270359

**Authors:** Samantha K Davis, Kevin John Selva, Ester Lopez, Ebene R Haycroft, Wen Shi Lee, Adam K Wheatley, Jennifer A Juno, Amy Adair, Phillip Pymm, Samuel J Redmond, Nicholas A Gherardin, Dale I Godfrey, Wai-Hong Tham, Stephen J Kent, Amy W Chung

## Abstract

Following infection with SARS-CoV-2, virus-specific antibodies are generated which can both neutralise virions and clear infection via Fc effector functions. The importance of IgG antibodies for protection and control of SARS-CoV-2 has been extensively reported. In comparison, other antibody isotypes including IgA have been poorly characterized. Here we characterized plasma IgA from 41 early convalescent COVID-19 subjects for neutralisation and Fc effector functions. We find that convalescent plasma IgA from >60% of the cohort have the capacity to inhibit the interaction between wild-type RBD and ACE2. Furthermore, a third of the cohort induced stronger IgA-mediated inhibition of RBD binding to ACE2 than IgG, when tested at equivalent concentrations. Plasma IgA and IgG from the cohort, broadly recognize similar RBD epitopes and showed similar ability to inhibit ACE2 from binding 22 of 23 different prevalent RBD proteins with single amino acid mutations. Plasma IgA was largely incapable of mediating antibody-dependent phagocytosis in comparison to plasma IgG. Overall, convalescent plasma IgA contributes to neutralisation towards wild-type RBD and various RBD single mutants in most subjects, although this response is heterogeneous and less potent than IgG.

## Introduction

Severe acute respiratory syndrome coronavirus 2 (SARS-CoV-2), the causative agent of Coronavirus disease 2019 (COVID-19) has infected millions of people and caused over 5.6 million deaths globally since its discovery. SARS-CoV-2 trimeric spike protein consists of two domains; spike 1 (S1) and spike 2 (S2) (Wrapp et al., 2020). The receptor binding domain (RBD) within the S1 engages with angiotensin-converting enzyme 2 (ACE2) on human cells contributing to infection (Wrapp et al., 2020). Antibodies generated towards RBD following infection or vaccination can block engagement with ACE2 and neutralise SARS-CoV-2 (Collier et al., 2021; Lustig et al., 2021; Wheatley et al., 2021). Mutations within the RBD can generate variants that have improved transmissibility and can lead to escape of antibody immunity from vaccination or infection, thus potentially becoming variants of concern (VOC) (Dupont et al., 2021; Falsey et al., 2021; Rössler et al., 2021). Importantly, both neutralising and non-neutralising antibodies engage fragment crystallisable (Fc) receptors on immune cells (e.g. monocytes) to activate Fc effector functions to clear infection (Chan et al., 2021; Tauzin et al., 2021; Ullah et al., 2021; Winkler et al., 2021). This polyfunctional antibody response assists in protection and control of viral infections, such a SARS-CoV-2 (Natarajan, Crowley, et al., 2021a; Sadarangani et al., 2021).

SARS-CoV-2 infection and vaccine-induced plasma antibodies directed towards RBD have been widely reported to neutralise SARS-CoV-2 using the fragment antigen binding (Fab) portion (Devarasetti et al., 2021; Lopez et al., 2021; Wheatley et al., 2021). Convalescent plasma IgG and IgM have been heavily studied, with both isotypes playing a large role in the neutralising response to SARS-COV-2 wild-type (WT) and VOC’s (Gasser et al., 2021; Liu et al., 2020; Maeda et al., 2021). Limited studies suggest plasma IgA dominates neutralisation of SARS-CoV-2 WT during early infection and decreases into convalescence (Klingler et al., 2020; Sterlin et al., 2021). Importantly, some IgA monoclonal antibodies (mAb’s) (monomeric and dimeric) can neutralise SARS-CoV-2 WT very effectively (Ejemel et al., 2020; Wang et al., 2021). The importance of plasma IgA induced by infection with SARS-CoV-2 WT and their capacity to inhibit ACE2 binding to RBD mutants remains to be assessed.

Neutralising and non-neutralising antibodies, including IgG and IgA, can engage Fc receptors (FcγR and FcαR respectively) on immune cells and activate Fc effector functions to clear viral infection (Asokan et al., 2020; Blutt et al., 2012; Davis et al., 2020). Effector functions including complement activation, phagocytosis, and antibody dependent cellular cytotoxicity (ADCC) have been implicated in clearance and control of SARS-CoV-2 infection (Dufloo et al., 2021; Lee et al., 2021; Natarajan et al., 2021). Importantly, compromised Fc effector function and reduced neutralising potency have been linked to poorer disease outcomes in humans (Garcia-Beltran et al., 2021; Zohar et al., 2020). The necessity for a polyfunctional IgG antibody response has also been highlighted in animal models, where neutralising human IgG monoclonal antibody (mAb) therapy protects from SARS-CoV-2 infection when given as a prophylactic mAb but requires additional Fc functions for optimal protection when given as a therapeutic (Chan et al., 2021; Winkler et al., 2021). However, the importance of plasma IgA for optimal Fc effector function has yet to be characterised.

The functional capacity of polyclonal convalescent IgA to SARS-CoV-2 has yet to be characterised, especially to RBD mutations associated with VOC. Here we examined purified fractions of plasma and purified IgA and IgG, to investigate the contribution of these isotypes to the polyclonal convalescent antibody response to RBD and prevalent single amino acid RBD mutants.

## Results

### Robust antibody recognition and ACE2 binding inhibition by SARS-CoV-2 convalescent plasma

41 SARS-CoV-2 convalescent subjects (median age of 55; IQR 49-61) ∼41 days post symptom onset and 26 uninfected subjects (mean age of 54; IQR 24-60) donated plasma samples (sup. table 1.). To determine Spike-1 (S1) and RBD wild-type (RBDWT) antibody binding, convalescent and uninfected subject plasma were assessed for IgM, IgG, and IgA binding using a SARS-CoV-2 multiplex bead array (Selva et al., 2021b) (fig. 1, sup. fig. 1a-c). Most SARS-CoV-2 convalescent subjects generated IgM, IgG, and IgA antibodies to RBDWT (IgM: 73.15% positive, median MFI=54086; IgG: 100% positive, median MFI=45745; IgA 97.56% positive, median MFI=2495) (fig. 1a-c).

**Figure 1.**
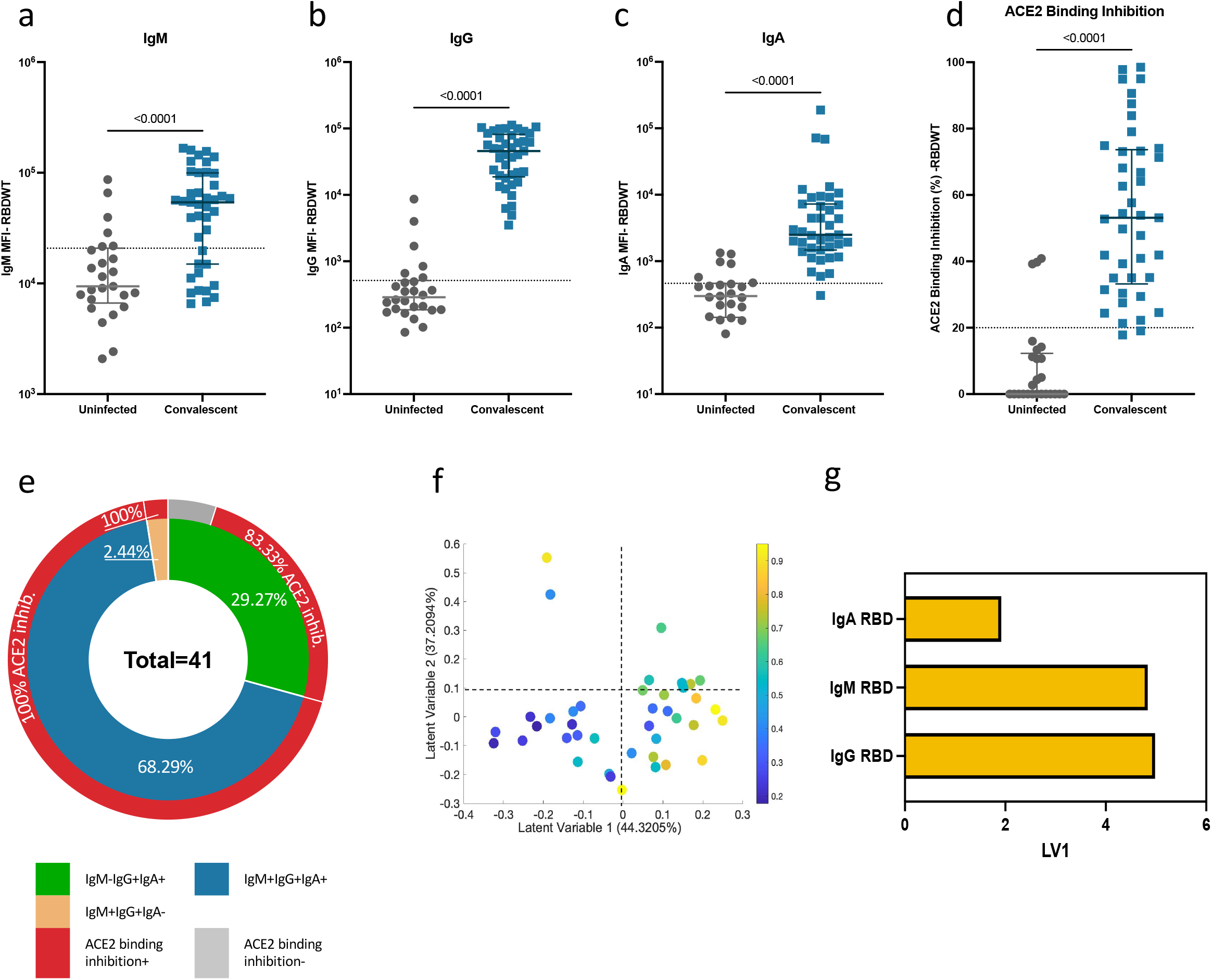
Convalescent plasma induced robust anti-SARS-CoV-2 RBDWT antibody isotypes levels that inhibit ACE2 from binding RBDWT. Convalescent (n-41) (blue) and uninfected control (n=26) (grey) plasma (final dilution 1:100) was assessed for IgM **(a)**, IgG **(b)** and IgA **(c)** antibody binding to SARS-CoV-2 RBDWT via multiplex. A positive threshold (grey dotted line) was defined as the 75^th^ percentile of antibody binding (MFI) for uninfected control plasma to respective antigens. **(d)** RBDWT ACE2 binding inhibition (%) of convalescent (blue) and uninfected control (grey) plasma (diluted 1:100). A positive threshold (grey dotted line) was defined as >20% ACE2 binding inhibition. **(e)** Pie chart outlining the percentage of subjects seropositive for anti-RBDWT antibody isotypes (IgM-IgG+IgA+ (green), IgM+IgG+IgA+ (blue), IgM+IgG+IgA-(yellow)) in the inner ring and the percentage of each seropositive subset with ACE2 binding inhibition in the outer red ring. Partial Least Squares Regression (PLSR) were conducted to determine the multivariate relationship between anti-RBD-specific isotype antibodies (IgG, IgA, and IgM) and % RBDWT-ACE2 binding inhibition (% ACE2 binding inhibition illustrated as a colour gradient legend on the right-yellow-strongest to dark blue-weakest). PLSR Scores **(f)** and loadings plot **(g)**. Percent variance for each latent variable (LV) in parentheses.

To examine the ability of subject plasma to neutralise SARS-CoV-2, we used a multiplex RBDWT-ACE2 binding inhibition assay, that has been demonstrated to highly correlate with gold standard live microneutralization assays (Lopez et al., 2021). Almost all convalescent subjects (95.12%) generated significant inhibition of ACE2 binding to RBDWT (median=73.23%, with ACE2 binding inhibition <20% considered to be below the limit of dection (Lopez et al., 2021)) (fig. 1d). Significant S1-ACE2 binding inhibition was also observed for convalescent subject plasma compared to uninfected subjects (sup. fig. 1d & e). Thus, most SARS-CoV-2 convalescent plasma induced antibodies that recognise RBDWT and have the capacity to inhibit the binding of RBD to ACE2, as previously reported (Lopez et al., 2021).

RBDWT antibody isotype titres (IgG, IgM and IgA) have previously been correlated to the level of plasma neutralisation or ACE2 binding inhibition (Gasser et al., 2021; Lopez et al., 2021; Sterlin et al., 2021). Here the cohort was broken down by isotype seropositivity and the ability to block ACE2 engagement with RBDWT to observe the surface level contribution of the isotypes to neutralisation in this cohort (fig. 1e). The majority (68.29%, 28 out of 41) of the convalescent cohort were seropositive for all 3 isotypes (RBDWT IgM, IgG and IgA) with 100% (28 out of 28) of these individuals being positive for ACE2 binding inhibition (fig. 1e). 29.27% (12 out of 41) of the cohort were seronegative for IgM but seropositive for IgG and IgA, with 83.33% (10 out of 12) of these individuals being positive for ACE2 binding inhibition (fig. 1e). A single individual (2.44% of the cohort) was seronegative for IgA but seropositive for IgM and IgG and could mediate ACE2 binding inhibition (fig. 1e). To investigate the combined contribution of anti-RBDWT antibody isotypes (IgG, IgM and IgA) to ACE2 binding inhibition, we performed partial least squares regression (PLSR) (fig. 1f-g). Not surprisingly, plasma samples with the highest ACE2 inhibition (yellow – highest, dark blue weakest ACE2 inhibition) predominantly cluster together across Latent Variable 1 (X-axis fig. 1f) and were strongly associated with anti-RBDWT antibody isotype titres, with IgG and IgM being the strongest correlates (fig. 1g). This was further confirmed by individual correlations with anti-RBDWT antibody isotypes (sup. fig. 2). Similar trends were observed for S1 (sup. fig. 1a-e). This data highlights that robust anti-RBDWT IgM, IgG and IgA response develops against SARS-CoV-2 and persist during early convalescence.

### IgA and IgG antibody depletion from convalescent plasma reduces ACE2 binding inhibition

IgG has been shown to play a critical role in plasma neutralisation to various viral infections, including SARS-CoV-2 (W. T. Lee et al., 2021; Lopez et al., 2021; Maeda et al., 2021). However, the contribution of plasma IgA to the convalescent SARS-CoV-2 neutralising antibody response, especially to emerging RBD mutants remains unclear (Sterlin et al., 2021; Verkerke et al., 2021; Wang et al., 2021). To define the contribution of IgG and IgA to the neutralising capacity of convalescent plasma, we depleted IgA from plasma (IgA-plasma) and also depleted plasma of both IgA and IgG (IgA- and IgG-depleted plasma) to assess the capacity of antibody depleted plasma fractions to inhibit RBDWT binding to ACE2 (fig. 2, sup. fig. 3). Successful IgG and IgA depletion was confirmed by a significant reduction in IgA and IgG binding to RBDWT (p<0.0001) and S1 (p<0.0001) for IgA- and IgA-/IgG-plasma (sup. fig. 3f-g, i-j). 73.2% (30 out of 41) of convalescent IgA-plasma samples were successfully depleted of IgA and had <30% loss of IgG or IgM (sup. fig. 3b).

**Figure 2.**
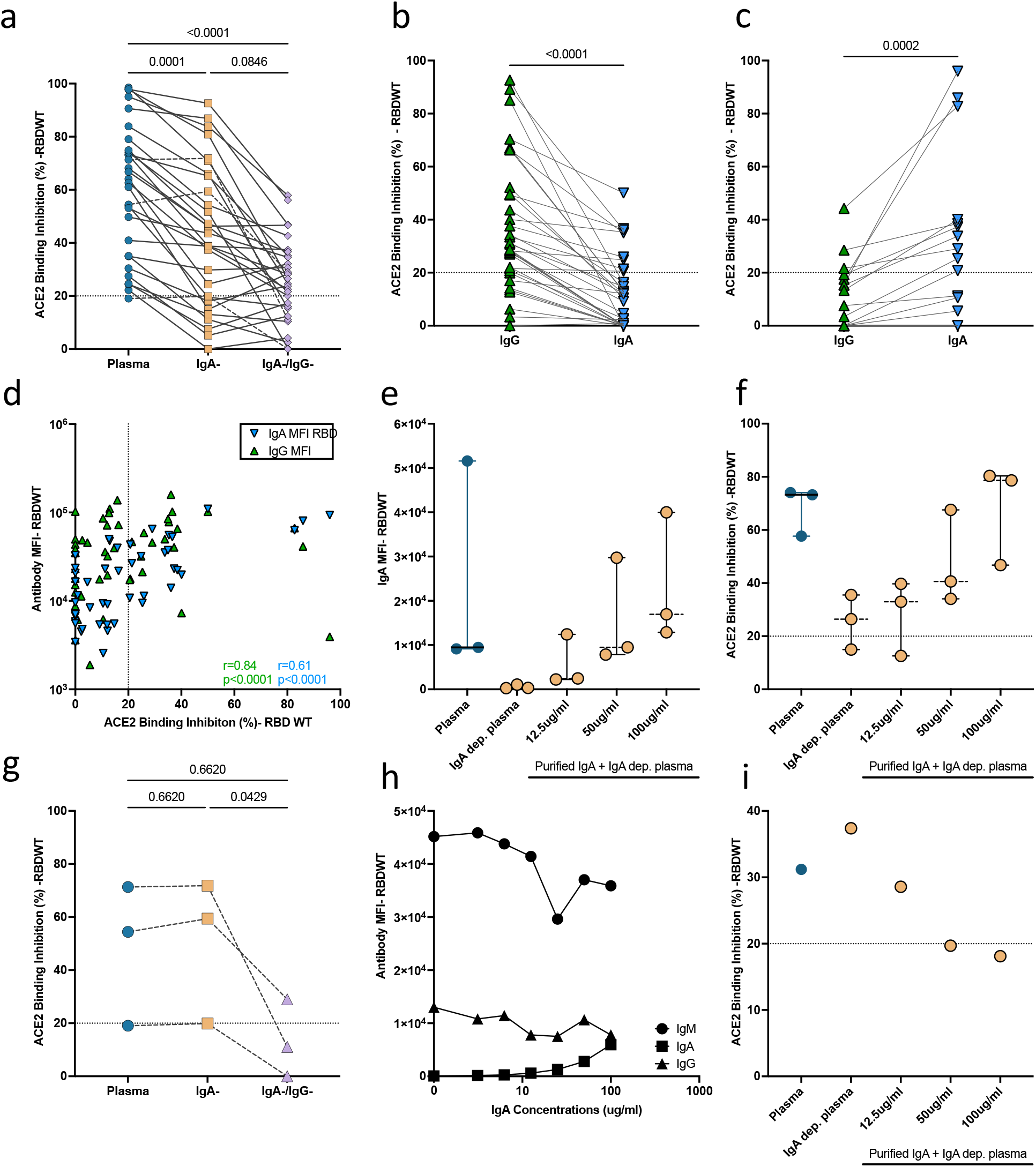
IgA from convalescent plasma induces variable levels of % RBD-ACE2 binding inhibition. SARS-CoV-2 RBDWT ACE2 binding inhibition (%) of **(a)** convalescent plasma (diluted 1:200; blue) and matched dilutions of IgA depleted (IgA-; yellow) and IgA plus IgG depleted (IgA-/IgG-; purple) (n=30) plasma fractions. Comparisons between plasma and depleted fractions was conducted using Friedman with Dunn’s multiple comparison test. (sup. Fig. 3 describes inclusion/exclusion criteria for successful antibody depletion); Comparison of matched purified IgG (green) and IgA (blue) RBDWT ACE2 binding inhibition tested at 100μg/ml total purified antibody separated as individuals where **(b)** purified IgG or **(c)** purified IgA mediated enhanced ACE2 binding inhibition. **(d)** Correlation (non-parametric Spearman test) between RBDWT-ACE2 binding inhibition (%) and anti-RBDWT antibody IgG and IgA isotype binding (MFI) tested at 100μg/ml total antibody. **(e)** RBDWT IgA binding (MFI) and **(f)** ACE2 binding inhibition (%) were assessed as increasing concentrations (yellow, 0, 12.5, 50 and 100μg/ml total antibody) of purified COVID+ IgA was spiked back into IgA depleted plasma (n=3). While **(g)** samples where IgA depletion resulted in an increase in RBDWT ACE2 binding inhibition **(h)** anti-RBDWT antibody IgG, IgA and IgM isotype binding (MFI) and **(i)** ACE2 binding inhibition (%) were assessed as increasing concentrations of purified IgA were spiked back into respective IgA-depleted plasma for this individual.

ACE2 binding inhibition of whole plasma at 1:100 was compared to matched dilutions of IgA- and IgA-/IgG-plasma to investigate the contribution of IgA and IgG to convalescent plasma neutralisation (fig. 2). IgA depletion reduced the capacity for convalescent plasma to mediate RBDWT-ACE2 binding inhibition by 22.14% (IgA depletion: median ACE2 inhibition=41.24%, IQR 19.19%-65.41%,) compared to matched complete convalescent plasma (median ACE2 inhibition=63.38%, IQR 35.03%-75.92%, p=0.0001) (fig. 2a). We also investigated the contribution of IgG to convalescent plasma neutralisation. IgG depletion (IgA-/IgG-plasma) further reduced RBDWT-ACE2 binding inhibition by 15.68% (median ACE2 inhibition= 25.56%, IQR 14.64%-33.96%, p=0.08) compared to IgA-plasma (fig. 2a). Similar trends were observed for S1 (sup. fig. 1f-h). This data suggests both IgA and IgG contributes to the neutralising capacity of convalescent plasma for most individuals in this cohort.

### The contribution of convalescent purified IgA to RBDWT-ACE2 binding inhibition is heterogenous

Although the above depletion studies suggest IgA contributes to convalescent plasma neutralisation in the majority of the cohort, a more definitive role of IgA or IgG in neutralisation could be revealed by specifically purifying these fractions and measuring antibody binding using a multiplex bead array. First, we investigated the capacity for purified IgG and IgA to mediate ACE2 binding inhibition to RBDWT at 100μg/ml total antibody. Purified convalescent IgG (median=27.01% IQR 13-37%-41.87%) and IgA (median=12.17% IQR 0-31.39) mediated increased RBDWT-ACE2 binding inhibition compared to purified IgA (median=12.57% IQR=5.633%-13.22%) and IgG (median=21.69% IQR=14.60-29.01) from uninfected donors, although this increase was not significant (IgA p=0.7810; IgG p=0.5665) (sup. fig. 4a-h). Not surprisingly a smaller proportion of individuals had detectable neutralization levels (>20% ACE2 inhibition; Purified IgA: 26.5% and Purified IgG: 59.18% of individuals, sup. fig 4c, f-h), than predicted by the IgA/IgG depletion experiments, likely due to the low concentration of purified IgA (100μg/ml) antibody available for use in these assays.

We next compared the ACE2 inhibitory capacity of matched purified IgG and IgA from the same individuals at equivalent total purified antibody concentrations (100μg/ml). Overall, within this cohort IgG mediated significantly higher ACE2 binding inhibition compared to IgA (p=0.0045) (sup. fig. 4h). However, examining this cohort in greater detail, while 69.39% (34 out of 49) individuals had lower IgA mediated ACE2 binding inhibition (fig. 2b, sup. fig 4h), interestingly, 30.1% (15 out of 49) of individuals mediated stronger ACE2 binding inhibition via IgA instead of IgG (fig. 2c). Furthermore, the top 3 purified IgA neutralisers had comparable IgA mediated neutralisation when compared to the top 3 purified IgG neutralisers (fig. 2b & c). Similar trends were observed for S1 antibody binding and ACE2 binding inhibition for purified convalescent IgG and IgA (sup. fig. 4a-c). We also demonstrate that the level of RBD-specific antibody binding correlated with ACE2 binding inhibition for both purified IgA (r=0.61, p<0.0001) and IgG (r=0.84, p<0.0001) (fig. 2d), suggesting that both IgG and IgA have the potential to neutralise SARS-CoV-2, however this is largely dependent on anti-RBDWT antibody titre/ concentration.

To confirm the importance of antibody titre for an individual’s ability to facilitate IgA mediated ACE2 binding inhibition, increasing concentrations of purified IgA (12.5, 50 and 100μg/ml total antibody) were spiked back into IgA-plasma from individuals with IgA mediated ACE2 binding inhibition (>20%) where sufficient sample was available (n=3) (fig. 2e). RBDWT-ACE2 binding inhibition of IgA-plasma increased with increasing concentrations of IgA, with a 2.98-fold increase in RBDWT-ACE2 binding inhibition compared to IgA-plasma (median ACE2 inhibition=26.43% IQR=14.98%-35.50%) with addition of 100μg/ml purified IgA (median ACE2 inhibition=40.61% IQR=34.05%-67.55%) (fig. 2f).

Intriguingly, upon further examination, we identified a small subset of individuals (n=3), where depletion of IgA resulted in a small increase in ACE2 inhibition (fig. 2g) (plasma: median ACE2 inhbition= 54.41%, IgA depleted plasma: median ACE2 inhbition= 59.41%). We selected the individual with the strongest level of RBDWT-ACE2 binding inhibition (CP30), and purified IgA was spiked back into IgA depleted plasma at increasing concentrations for this individual. IgM binding decreased (0.2-fold reduction, fig. 2h), whereas RBDWT-ACE2 binding inhibition (0.5-fold increase, fig. 2i) surprisingly increased by the addition of 100μg/ml purified IgA. These results demonstrate that in rare individuals, at higher titers, IgA may be able to block RBD-specific IgM/IgG from binding, thus inhibiting ACE2 inhibition by other isotypes. This inhibition is likely through IgA binding to non-neutralising epitope(s), thus blocking the binding of other neutralising antibodies isotypes.

### Antibody binding profiles of SARS-CoV-2 convalescent purified IgG and IgA to RBD single mutants

SARS-CoV-2 variants are rapidly emerging, that include amino acid mutations within the RBD, some of which confer increased ACE2 affinity and antibody escape (Harvey et al., 2021; Verma & Subbarao, 2021). The capacity for IgG mAbs, convalescent plasma and plasma IgG to recognise different RBD mutations has been widely characterised, however, the capacity of convalescent purified IgA to recognise different RBD mutations is yet to be reported (Lopez et al., 2021; Maeda et al., 2021). To explore the impact of RBD mutations on IgA and IgG driven ACE2 binding inhibition, plasma, IgA-depleted plasma and IgA-/IgG-depleted plasma were assessed for ACE2 binding inhibition to a panel of 23 RBD single amino acid mutants, using a previously published competitive RBD-ACE2 inhibition multiplex assay (Lopez et al., 2021). As expected, weaker whole plasma ACE2 inhibition was observed for a number of RBD single amino acid mutations including E484K (present in Beta and Gamma VOC) and N501Y (present in Beta, Gamma and Omicron) as compared to RBD-WT, as previously described (Lopez et al., 2021). Depletion of IgA and IgG also reduced RBD-ACE2 inhibition levels across all RBD variants, suggesting that IgA and IgG contribute to the recognition of RBD variants (fig. 3a, sup. fig. 5a).

**Figure 3.**
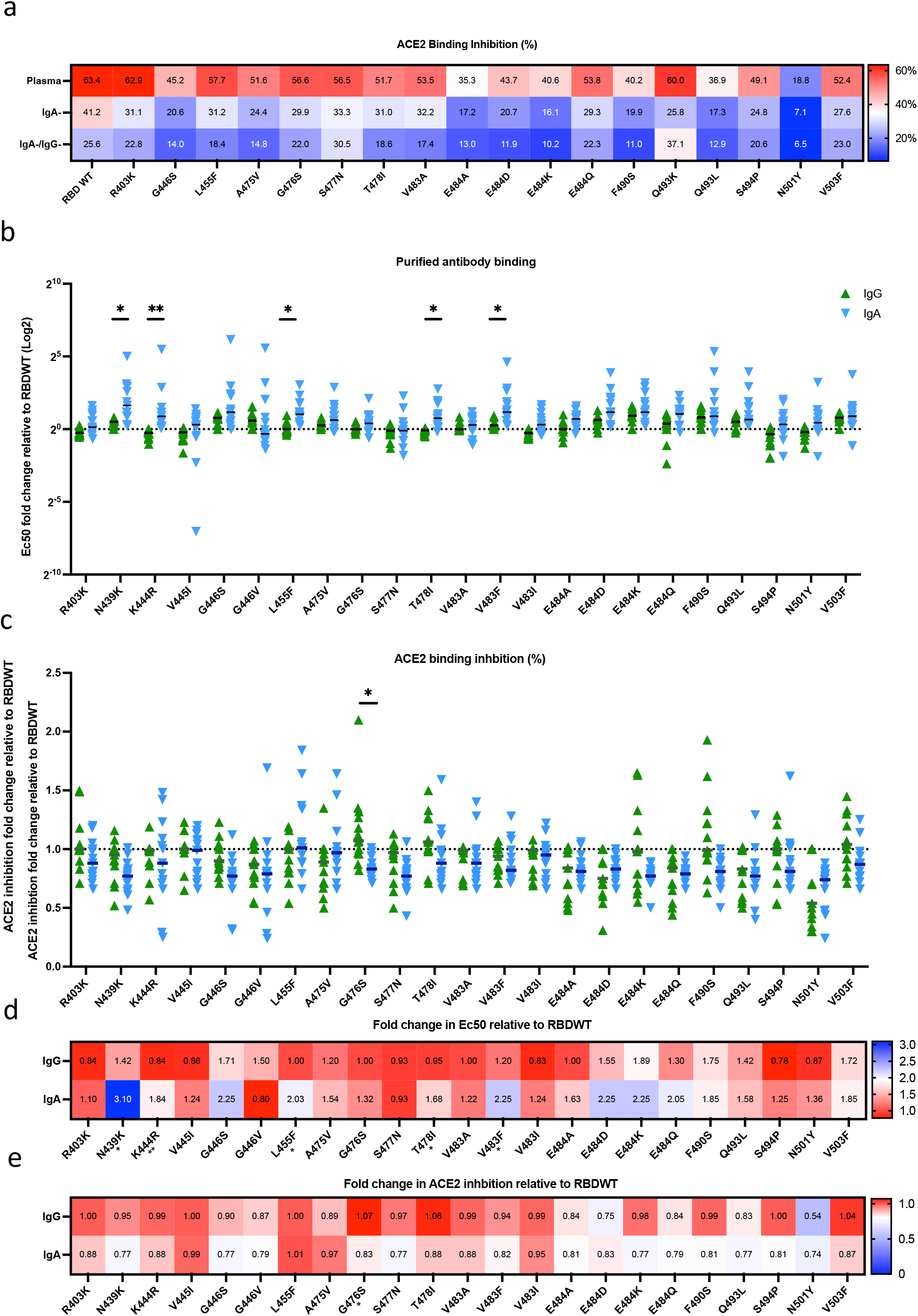
Similar antibody binding and ACE2 binding inhibition of convalescent purified IgA to RBD single mutants compared to IgG. **(a)** Heatmap of the mean ACE2 binding inhibition (%) for RBDWT and 18 RBD single mutants of matched complete plasma, IgA-depleted, and IgA-/IgG-depleted plasma (n=30). Purified IgA (blue) and IgG (green) **(b)** fold change in antibody EC_50_ binding relative to RBDWT for 23 different RBD single mutants of subjects (n=13) **(c)** Fold change in RBD-ACE2 binding inhibition relative to RBDWT for 23 different RBD single mutants of subjects for the same samples (n=13). Statistical analyses were performed with the Paired t test adjusting for multiple comparisons using the Holm-Šídák method (*=p<0.05, **=p<0.01). The median fold change in **(d)** antibody EC_50_ binding and **(e)** % RBD-ACE2 binding inhibition relative to RBDWT for each RBD mutant is summarized as heatmaps.

To further characterise the IgA and IgG-driven ACE2 binding inhibition to RBD mutations, individuals who mediated stronger IgA-mediated RBDWT-ACE2 binding inhibition (median: 28.31%, n=13) were characterised further for their matched purified IgG and IgA binding and ACE2 binding inhibition to 23 RBD single amino acid mutations (fig. 3b-e). First, antibody binding relative to RBDWT for 23 RBD single mutants was calculated using EC_50_ values obtained from the normalised antibody binding MFI for matched convalescent purified IgA and IgG samples (titrations up to 100μg/ml) (fig. 3b & d, sup. fig. 5b). Importantly, for 18 of the 23 mutants (78.26%) there was no significant difference in antibody binding for purified IgG and IgA (fig. 3b & d). Overall, IgG trended towards better antibody binding (lower EC_50_) compared to IgA, with 5 RBD mutants (L455F (p= 0.0153), N439K (p=0.0153), T478I (p=0.0107), K444R (p=0.0056) and V483F (p=0.032), all p-values adjusted for multiple comparison) having significantly greater IgG recognition compared to IgA using multiple comparisons (fig. 3b & d). Interestingly, the RBD mutant G446V showed a trend towards (adjusted p=0.0956) better IgA recognition relative to RBDWT (median EC_50_=157.7 IQR=68.86-180.9) compared to IgG (median EC_50_=246.5 IQR=108.9-400) (fig. 3b & d, sup. fig. 5b). Notably, we observed a larger range in fold change for individual IgA binding (median range of fold change across all variants=7.017) relative to RBDWT for the 23 RBD mutants compared to IgG (median range of fold change across all variants=0.64) (fig. 3b). This data suggests IgG and IgA can recognise RBD mutations similarly.

We next investigated if differences in antibody recognition by IgG and IgA translated to a difference in ACE2 binding inhibition. We assessed the ACE2 binding inhibition capacity of purified IgA and IgG for the same IgA neutralisers (n=13) to 23 RBD single mutants at a single concentration (100μg/ml total antibody) using the previously described multiplex ACE2-inhibition array (fig. 3c & e). Purified IgG had significantly increased ACE2 binding inhibition relative to RBDWT compared to purified IgA for 1 RBD mutation (G476S) (p=0.011) (fig. 3c). Purified IgG had enhanced recognition of G476S compared to purified IgA (not significant), which may suggest that IgG and IgA may recognize different epitopes for this mutant (fig. 3b, sup. fig. 5c). Importantly, for 22 of the 23 mutants (95.65%) there were marginal differences in ACE2 binding inhibition for purified IgG and IgA, suggesting that IgG and IgA can neutralise most RBD mutants similarly. Taken together, this data suggests that purified convalescent IgA and IgG have similar capacities to recognise most RBD mutations assayed here. Where significant differences were observed in IgG and IgA antibody recognition, ACE2 binding inhibition or neutralisation was marginally impacted.

### SARS-CoV-2 specific IgG mediates Fc effector function of convalescent plasma

Antibody Fc portions can engage with FcyR on monocytes to mediate antibody dependent phagocytosis (ADP). Anti-SARS-CoV-2 IgG mAb’s and plasma IgG has been reported to induce robust ADCP responses, however, the Fc functional role of plasma IgA in SARS-CoV-2 is yet to be characterised. Here, we investigated the capacity of convalescent plasma to induce ADP using a previously described SARS-CoV-2 ADP bead assay (W. S. Lee et al., 2021). We measured the antibody-mediated uptake of spike trimer (S)-conjugated fluorescent beads by THP-1 monocytes with a subset of convalescent (n=18) and uninfected control plasma (n=12) (fig. 4a). All convalescent subjects had higher ADP (median phagocytic score=9.31 × 10^3^ IQR= 7.14 × 10^3^ – 11.20 × 10^3^), compared to uninfected subjects (median phagocytic score=0.11 × 10^3^ IQR=0 × 10^3^ −0.45 × 10^3^, p<0.0001) (fig. 4a). To determine the contribution of IgA and IgG to the ADP response against SARS-CoV-2, we measured ADP activity mediated by IgA- and IgA-/IgG-plasma. (fig. 4b). Depletion of IgA had no effect on the ADP capacity of the convalescent plasma (convalescent plasma: median phagocytic score=9.31 × 10^3^; IgA-plasma: median phagocytic score=10.01 × 10^3^ IQR= 5.91 × 10^3^ −12.98 × 10^3^, p>0.999) (fig. 4b). Conversely, depletion of IgG (IgA-/IgG-plasma) (median=2.20 × 10^3^ IQR= 1.36 × 10^3^ −3.11 × 10^3^) resulted in a median loss of 76.37% ADP activity compared to convalescent plasma (p<0.0001) (fig. 4b). This data highlights the importance of the IgG antibody isotype in the Fc functional capacity of SARS-CoV-2 convalescent plasma.

**Figure 4.**
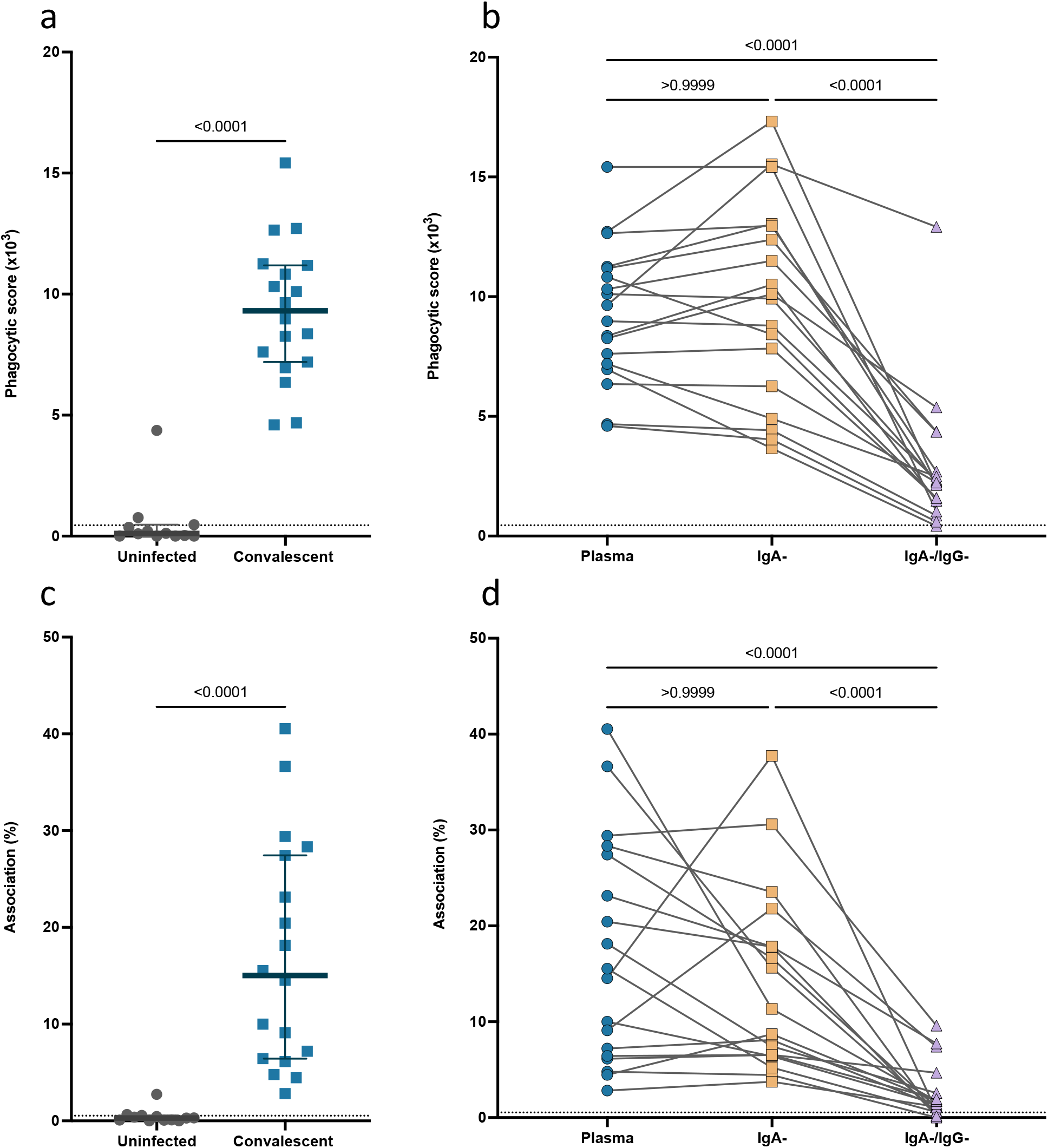
Depletion of IgG but not IgA reduces the Fc functional capacity of convalescent plasma. **(a)** Antibody dependent phagocytosis (ADP) of S-conjugated beads for a subset of convalescent subject plasma (blue) (n=18) and uninfected control plasma (grey) (n=12) (diluted 1:900). **(b)** Comparison of ADP of S-conjugated beads from matched convalescent subject plasma (blue), IgA depleted plasma (IgA-; yellow) and IgA and IgG depleted plasma (IgA-/IgG-; purple) (n=18). **(c)** Fcy Receptor (FcyR) dependent association of THP-1 cells with Ramos S-orange cells mediated by convalescent plasma (blue) and uninfected control plasma (grey) (diluted 1:100). **(d)** FcyR dependent association of THP-1 cells with Ramos S-orange cells from matched convalescent subject plasma (blue), IgA depleted plasma (IgA-; yellow) and IgA and IgG depleted plasma (IgA-/IgG-; purple) (n=18). Grey dotted lines denote the negative threshold for cell association (<2%) and ADP (3 × 10_3_) calculated as the mean response of negative subject plasma plus 2 standard deviations. Statistical analyses were performed with the Kruskal-Wallis test followed by Dunn’s multiple comparisons test.

In addition to uptake of antibody opsonised virions, antibodies can also mediate cell association and trogocytosis of infected cells expressing SARS-CoV-2 spike. Using an established cell association assay (W. S. Lee et al., 2021), we measured antibody mediated association of THP-1 monocytes with cells expressing SARS-CoV-2 spike (Ramos S-orange cells) following incubation with convalescent (n=18) and uninfected subject plasma (n=12) (fig.4c). Antibody mediated association was detected in all convalescent subjects (median association=15.04% association IQR= 6.63%-27.66%) when compared to uninfected subjects (median association=0.3% association IQR= 0.08%-0.53%, p<0.0001) (fig. 4c). We then investigated the contribution of IgA and IgG to cell association/ trogocytosis through depletion of IgA (IgA-plasma) and IgG (IgA-/IgG-depleted plasma) from convalescent plasma (fig. 4d). Depletion of IgA (median association=10.02% IQR=6.50%-18.83%) did not significantly alter the cell association mediated by convalescent plasma (median association=15.04% association IQR= 6.63%-27.66%, p>0.99) (fig.4d). However, depletion of IgG (IgA-/IgG-plasma) (median association=1.33% association IQR=0.61%-3.10%) resulted in a median loss of 90.53% cell association relative to convalescent plasma (median association=15.04% association IQR= 6.63%-27.66%, p<0.0001) (fig. 4d). This data further supports the role of IgG convalescent plasma for an effective Fc functional response to SARS-CoV-2.

## Discussion

A strong correlate of protection for most viral vaccines, including SARS-CoV-2 vaccines, are neutralising antibodies (Khoury et al., 2021; Plotkin, 2010). IgM and IgG have been widely implicated in effective neutralisation of SARS-CoV-2 (Gasser et al., 2021; Lopez et al., 2021; Natarajan, Xu, et al., 2021; Wheatley et al., 2021). However, throughout the COVID-19 pandemic, the study of anti-SARS-CoV-2 IgA responses have been relatively neglected (Verkerke et al., 2021). The plasma IgA response to SARS-CoV-2 peaks during acute infection but is relatively transient in nature, dominating the acute plasma neutralising response with a 2 phased decay in antibody binding to spike (half-life of 42 days in the first 60 days and >1000 days from 60-160 days post symptom onset) (Sterlin et al., 2021; Wheatley et al., 2021). Using our surrogate neutralisation assay, we show that anti-RBD plasma IgA present during early convalescence contributes to the neutralising capacity, in addition to IgG and IgM isotypes, for many individuals. As observed by Gasser et al., (2021), we also found decreased neutralisation when IgA was depleted from convalescent plasma, supporting IgA’s contribution to SARS-CoV-2 neutralisation. Furthermore, for the first time we show that IgA depletion and IgG depletion also reduces neutralisation for several RBD mutations.

We observed IgA-associated and IgA-independent neutralising antibody responses within this convalescent cohort. An IgA-independent neutralising response was observed in 30.3% of individuals with no change in ACE2 binding inhibition when IgA was depleted. While 69.7% of this cohort showed a significant reduction in neutralisation when IgA was depleted, suggesting IgA contributes to neutralization within these individuals. Furthermore, similar to Wang et al., (2021) a subset of convalescent individuals mediated superior neutralisation with IgA when compared to IgG. However, purified IgG mediated higher RBD-ACE2 inhibition compared to purified IgA for most convalescent donors. While IgG and IgA contribute to convalescent plasma neutralisation, depletion of IgG and IgA did not abolish neutralisation in many individuals, supporting the role of IgM in the convalescent plasma neutralising response to SARS-CoV-2 (Gasser et al., 2021; Natarajan, Xu, et al., 2021). Our data further supports the finding that IgA is capable of neutralising SARS-CoV-2 WT, although this response is heterogenous with in this cohort, likely due to heterogeneity in IgA titres against RBD.

This was confirmed by spiking increasing concentrations of purified IgA back into IgA-plasma, resulting in increased capacity for plasma to mediate SARS-CoV-2 neutralisation. IgA has been widely reported to be broadly cross reactive towards human coronaviruses, particularly in the elderly (Selva et al., 2021a). Therefore, it is also possible that pre-existing IgA+ memory B cells for human coronaviruses may recognise SARS-CoV-2 RBD with higher affinity and undergo somatic hypermutation to create a potent and robust IgA response in some individuals (Wec et al., 2020). Another possible explanation for the variability between individuals could be preferential class switching to IgG or IgA due to the cytokine environment and/or host genetics. Although, cytokine analysis or sequencing of gamma and alpha genes were not in the scope of this project, the cytokine profiles induced by SARS-CoV-2 vary with disease severity (Ghazavi et al., 2021) and may impact class switching(Cerutti, 2008; Stavnezer, 1996).

Intriguingly, we also observed a single individual which showed decreased neutralisation and IgM binding to RBDWT when increasing concentrations of purified IgA were spiked back into IgA-depleted plasma. This suggests IgA in rare cases may block RBD-specific IgM antibodies from binding and neutralising SARS-CoV-2. Similarly, HIV-1 vaccine induced IgA antibodies have previously been reported to block binding of IgG to a protective epitope and prevent neutralisation and effector functions (Tomaras et al., 2013). However, in most instances anti-SARS-CoV-2 convalescent IgA has the capacity to neutralise wild-type SARS-CoV-2 when sufficient titres of IgA are present.

As new variants of concern emerge consisting of a constellation of single amino acid substitutions within the RBD, it is important to understand antibody isotype (IgG and IgA) recognition and neutralising capacity to these single site mutations. Single amino acid substitutions can alter the stability and potentially the structure of the RBD, thus may reveal or hide epitopes from different isotypes and/or subclasses of antibodies (Chowdhury et al., 2020; Verma & Subbarao, 2021). For individuals with IgA-skewed neutralisation, we compared the capacity for polyclonal purified convalescent IgA to recognise and neutralise single amino acid RBD mutants compared to IgG. We observed minor differences in purified antibody isotype binding to various RBD single mutants, despite the small sample sizes and the polyclonal nature of these convalescent responses. A trend toward preferential IgG binding to many mutants was observed, with significantly increased IgG binding to 5 mutations. Interestingly, we observed a trend towards increased IgA binding to a single RBD mutation; G446V, which is predicted to reduce RBD stability and increase ACE2 affinity (Verma & Subbarao, 2021). The Fc portion of the antibody can affect the fine epitope specificity (binding affinity/epitope recognition) of the Fab region which could explain slight variation in IgG and IgA recognition of RBD mutants (Casadevall & Janda, 2012; Ejemel et al., 2020; Janda et al., 2012; Tudor et al., 2012). Furthermore, increased flexibility of the hinge of IgA1, the predominant subclass of IgA found in blood, may allow for increased accessibility to certain epitopes of Spike and RBD compared to IgG, such as the G446V mutation (Davis et al., 2020; Ejemel et al., 2020). Overall, our results show that convalescent IgG and IgA broadly recognise similar RBD epitopes with only minor differences in the ability to neutralise RBD mutants.

In addition to antibody neutralisation, Fc effector functions such as phagocytosis, are important for control and clearance of SARS-CoV-2 infection (Butler et al., 2021; Chan et al., 2021; Winkler et al., 2021). Similar to previous studies, we show that convalescent plasma can mediate ADP (Butler et al., 2021, W. S. Lee et al., 2021). Interestingly, IgA depletion from convalescent plasma did not impact Fc effector functions by THP-1 monocytes. THP-1 cells express relatively low amounts of FcαR, and thus we may not capture the functional potential of IgA the has been alluded to by Butler et al., (2021) who suggested convalescent IgA contributes to the functional capacity of convalescent plasma through regression analysis. Importantly, depletion of IgG abolished Fc effector functions in our assays, further reinforcing the importance of IgG in the polyfunctional (neutralising and Fc effector function) antibody response of convalescent plasma.

Overall, we find that convalescent plasma IgA recognises RBDWT and an array of RBD mutants and has the capacity to block ACE2 engagement with RBD in a comparable manner to IgG, when sufficient IgA titers are induced. Furthermore, the convalescent IgA neutralising response is highly heterogenous between individuals, with a third of the cohort inducing stronger IgA-mediated inhibition of RBD engagement with ACE2 than IgG, when tested at equivalent concentrations. Dissecting the IgA response in the context of vaccination and to variants of concern is essential to further understand the importance of IgA in a protective polyclonal antibody response.

## Materials and Methods

### Ethics statement

The study protocols were approved by the University of Melbourne Human Research Ethics Committee (#2056689) and all associated procedures were carried out in accordance with the approved guidelines. All participants provided written informed consent in accordance with the Declaration of Helsinki.

### Human Subjects

Participants who had recovered from COVD-19 during the first wave of the pandemic in Melbourne, Australia, March-May 2020, were recruited as previously described (Wheatley et al., 2021). Convalescent subjects were confirmed to have had COVID-19 by returning a positive PCR test during early infection or had clear exposure to SARS-CoV-2 and tested positive for SARS-CoV-2 serology (both Spike trimer and RBD) confirming prior exposure as previously reported (Juno et al., 2020). Uninfected controls with no COVID-19 symptoms were also recruited during the first wave of COVID-19 and were confirmed as seronegative. Whole blood was collected with sodium heparin. The plasma fraction was then collected and stored at −80°C. Cohort characteristics for convalescent and healthy controls are outlined in Table S1.

### Human IgG and IgA ELISA

Purified IgG and IgA concentrations were quantified using human anti-IgG kit (cat# 3850-1AD-6 Mabtech) or anti-IgA kit (cat# 3860-1AD-6 Mabtech) respectively as per manufacturer’s instructions. Briefly, anti-IgG or IgA capture antibody was coated on Maxisorb 96 well plates (Nunc) overnight at 4°C. Plates were washed with PBS containing 0.05% Tween20 (PBST) and blocked with 1% BSA/PBST for 2 h. The plates were washed, and purified IgG or IgA antibodies were titrated 2-fold from 1:20,000 for a minimum of 4 points. To check for IgG contamination, purified IgA antibodies were tested at 1:1,000 and 1:2,000 dilutions. Similarly purified IgG antibodies were tested at 1:1,000 and 1:2,000 to check for IgA contamination. Antibodies and respective standards were incubated for 2h at RT before being washed. The secondary-ALP conjugated antibody was added to each well and incubated at RT for 1 h. The plate was washed, and the substrate p-nitrophenyl-phosphate (pNPP) was added and left to develop (∼30mins). The optical density at 405nm was read using Thermo Fisher Multiskan Ascent plate reader. Purified IgA and IgG was confirmed to be free of contamination from the other isotype if their OD was less than the background no antibody OD plus 2 standard deviations.

### Antibody purification and depletion

IgA purification and depletion from plasma samples were performed via affinity chromatography using peptide M agarose following manufacturer’s instructions (sup. fig. 3a). Briefly, peptide M agarose (Jomar) was added to 1ml filter columns (thermofisher) and washed 3 times with PBS. 300ul of plasma was incubated with peptide M columns for 45 minutes on an orbital at room temperature. Depleted IgA plasma fractions were collected by spinning at 1000g for 1 min after incubation. Purified IgA was eluted with low pH (pH2.8) by IgG Elution Buffer (Thermofisher). Elution was neutralised using Tris M pH 8.0 (Life Sciences).

IgG was purified from 100ul of IgA depleted (IgA-) plasma using 96 well Protein G HP MultiTrap (GE Healthcare) following manufacturer’s instructions (sup. fig. 3a & b). Purified IgA was also passed through the MultiTrap to remove any IgG contamination. Briefly, IgA-plasma and purified IgA was diluted 1:1 in antibody binding buffer and added to the MultiTrap. Samples were incubated for 30 mins at RT while shaking. The plate was centrifuged, IgA and IgG (IgA-/IgG-) depleted plasma and purified IgA was collected. The plate was washed with antibody binding buffer before elution of IgG with 200ul of elution buffer (GE Healthcare). Purified IgG was collected via centrifugation at 200xg for 2 mins and neutralised to pH 7 using neutralisation buffer (GE Healthcare). Elution was performed 3 times and purified IgG, IgA- and IgA-/IgG-plasma was buffer exchanged into PBS and concentrated to original starting volume.

### SARS-CoV-2 bead-based multiplex assay

The SARS-CoV-2 specific antibody isotypes (IgM, IgG and IgA1) were assessed using a multiplex assay as previously described (Selva et al., 2021). Briefly, bead mixture containing 700 beads per bead region and diluted plasma or purified antibody, was added to each well in a black clear bottom 384 well plate. Phycoerythrin (PE)-conjugated mouse anti-human pan-IgG and IgA1 (Southern Biotech) (1.3μg/ml) were added to detect SARS-CoV-2 specific antibodies. For IgM detection, biotinylated mouse anti-human IgM (mAb MT22; MabTech) was added at 1.3 μg/ml. Following incubation, streptavidin R-Phycoerythrin conjugate (SAPE, Invitrogen) at 1 μg/ml was added. The plate was read via the FlexMap 3D and binding of PE-detectors was measured to calculate the median fluorescence intensity (MFI). Background was corrected for by subtracting the MFI of BSA-blocked beads for each well. Titrations of pooled convalescent plasma and an anti-SARS-CoV-2 RBD neutralising human IgG1 antibody (SAD-S35, ACRO Biosystems, USA) were included as positive controls and uninfected subject plasma was included as negative controls. A single dilution was used for plasma, depleted plasma and EC_50_’s were calculated for purified IgG and IgA where appropriate using normalised antibody binding.

### RBD-ACE2 binding inhibition multiplex bead-based assay

RBD-ACE2 binding inhibition assay was performed as previously described (Lopez et al., 2021). Briefly, an array of SARS-CoV-2 antigens including S1 (Sino Biological), RBD wild-type (WT) (B; wild-type, Wuhan) and 18 RBD single mutants used in figure 3a with 5 additional RBD mutants were included in figure 3b-e, in a bead suspension containing 700 beads per bead region were added to each well (20ul), with biotinylated Avitag-ACE2 (kindly provided by Dale Godfrey, Nicholas Gherardin and Samuel Redmond) at a final concentration of 12.5μg/ml per well, and dilutions of plasma or purified antibodies were added to 384-well plates. Biotinylated Avitag-ACE2 was detected using SAPE at 4μg/ml followed by PE-Biotin amplifier (Thermo Fisher) at 10μg/ml. Plates were washed and acquired on a FlexMap 3D (Luminex). Anti-SARS-CoV-2 RBD neutralising human IgG1 antibody (SAD-S35, ACRO Biosystems, USA) was included as a positive control, in addition to COVID-19 negative plasma and buffer only negative controls.

The MFI of bound ACE2 was measured after background subtraction of no ACE2 controls. Maximal ACE2 binding MFI was determined by ACE2 only controls. % ACE2 binding inhibition was calculated as 100%−(% ACE2 binding MFI per sample/Maximal ACE2 binding).

### Matching dilutions to account for loss of antibody during depletion process

Anti-RBDWT IgM (sup. fig. 3c) and IgG (sup. fig. 3d) binding was also assessed for plasma, IgA- and IgA-/IgG-depleted plasma via multiplex. During the depletion process, a loss of 48.88% of IgG (sup. fig. 3c, median MFI=21150, p=0.0160) and 36.95% of IgM (sup. fig. 3d, median MFI=26544, p=0.0002) between medians was observed following the depletion of IgA (IgA-plasma) and IgG (IgA-/IgG-plasma) respectively compared to plasma (median IgG MFI=41377, median IgM MFI=42101) (sup. fig. 3c-d). To ensure fair comparisons between plasma and depleted plasma fractions, plasma and depleted plasma samples were titrated, and anti-RBDWT IgG and IgM binding MFI was determined via multiplex (process outlined in sup. fig. 3e). IgA- and IgA-/IgG-depleted plasma dilutions were chosen by matching IgG or IgM MFI respectively to plasma IgG or IgM binding MFI at a dilution of 1:100. IgA-plasma and IgA-/IgG-depleted plasma with final dilutions outlined in sup. table 2. These dilutions were used for all multiplex assays. Depleted samples with >30% loss in IgG or IgM following matching of dilutions were excluded from this study (sup. fig. 3b).

### Quality control testing of antibody depleted plasma

Successful depletion IgG and IgA was confirmed for matched dilutions via IgG SARS-CoV-2 RBDWT multiplex (sup. fig. 3f-k, sup. table 2). This method detected the remaining antigen specific IgG or IgA with high sensitivity which allowed for application of a stringent threshold for exclusion of samples with un-successful IgG or IgA depletion. Anti-RBDWT IgG or IgA binding MFI of depleted plasma was compared to full plasma for each subject and the percentage reduction in IgG and IgA binding was calculated Sufficient depletion was defined as >70% depletion of IgG or IgA (i.e. >70% reduction in anti-RBDWT IgG MFI compared to plasma IgG MFI) (sup. fig. 3f-k). Plasma, IgA-plasma and IgA-/IgG-depleted plasma samples with <70% IgA or IgG depletion in were all excluded from this study (sup. fig. 3b). Only subjects with all three plasma fractions (plasma, IgA- and IgA-/IgG depleted) were included in final analysis (n=30) (sup. fig. 3b).

### IgA spiking assay

Using RBDWT-ACE2 binding inhibition assay and the SARS-CoV-2 multiplex assay, purified IgA was spiked back into IgA-plasma up to 100μg/ml of IgA (12.5, 50 and 100μg/ml) as proof of concept that IgA contributes to ACE2 binding inhibition of convalescent plasma in a dose dependent manor.

### Fc effector functional assays

#### Cell culturing

THP-1 monocytes (ATCC) and Ramos cells expressing mOranage2 SARS-CoV-2 spike trimer (Ramos S-orange cells) (W. S. Lee et al., 2021) were cultured in RPMI 1640 with 10% FCS (RF10) under recommended cell culture conditions (37°C with 5% carbon dioxide (CO_2_)). THP-1 monocytes and Ramos S-orange cells were maintained below a cell density of 0.3 × 10^4^ and 1.0 × 10^4^ respectively. Flow cytometry was used to confirm stable expression of FcγRII (CD32), FcγRI (CD64) and FcαR (CD89) on THP-1 monocytes and stable expression of SARS-CoV-2 spike trimer (S-trimer) and expression of mOrange2 for Ramos S-orange cells. Cell viability was determined using trypan blue exclusion and morphology confirmed using light microscopy prior to assay preparation.

#### Antibody dependent bead-based phagocytosis assay

Antibody dependent bead-based phagocytosis assay was used as previously described (W. S. Lee et al., 2021). Briefly, SARS-CoV-2 spike trimer protein (S-trimer) (supplied by Adam Wheatly, The Peter Doherty Institute) was biotinylated and coupled to 1μm fluorescent NeutrAvidin Fluospheres (beads) (Invitrogen) overnight at 4°C. S-trimer coated beads were washed and diluted 1:100 in 2% BSA/PBS. 10ul of diluted S-trimer coated beads were incubated with plasma diluted 1:100 for 2 hours at 37°C in a 96 well U bottom cell culture plate. 10^4^ THP-1 monocytes were added to opsonized beads and incubated for 16 hours under cell culturing conditions. THP-1 monocytes were fixed, and cells were acquired by flow cytometry on a BD LSR Fortessa with a HTS (sup. fig. 6a for gating strategy). The data was analysed using FlowJo 10.7.1 and a phagocytosis score ([% bead positive cells × mean fluorescent intensity]/ 10^3^) was calculated as previously described Darrah, Patel et al. (2007).

#### THP-1 and Ramos S-orange cell association assay

A THP-1 and Ramos S-orange cell association was used as previously described (W. S. Lee et al., 2021). Briefly, THP-1 monocytes were stained with CellTrace™ Violet (CTV) (Life Technologies) as per manufacturer’s instructions. Concurrently, 10^4^ Ramos S-orange cells were added to each well in a 96-well V bottom cell culture plate. Plasma was diluted to 1:900 with the Ramos S-orange cells and incubated for 30 minutes under cell culture conditions. Opsonised Ramos S-orange cells were washed by centrifugation and 10^4^ of CTV stained THP-1 monocytes were added for a final 1:1 ratio of THP-1 monocytes to Ramos S-orange cells. Opsonised Ramos S-orange cells and THP-1 monocytes were incubated for 1 hour under cell culture conditions before fixation. Cells were acquired by flow cytometry using the BD LSR Fortessa with a high-throughput sampler attachment (HTS) and the data was analysed using FlowJo 10.7.1 (sup. fig. 6b for gating strategy). The percentage of Ramos S-orange cells associated with THP-1 monocytes (% association) was extracted.

### Data normalisation

For purified antibody binding to the RBD single mutants of subjects with IgA neutralisation (n=13), antibody MFI values were normalised to the maximum antibody binding (100%) (IgG and IgA) to each variant to account for differences in coupling efficiency for the RBD mutants. The normalised purified antibody binding was used to calculate EC_50_’s for purified IgG and IgA. EC_50_’s >400 were set to the threshold of 400.

### Data analysis

Traditional statistical analyses were performed with Graphpad Prism 9. See figure ledgends for details. Partial Least Squares Regression Analyses were conducted to determine multivariate relationships between immune features and continuous variables (e.g. ACE2 binding inhibition) using Matlab with the statistics and machine learning toolbox (Mathsworks) and PLS_Toolbox (Eigenvector).

## Data Availability

All data produced in the present study are available upon reasonable request to the authors

## Acknowledgments

This study was supported by the Medical Research Future Fund (MRFF) (W.-H.T., D.I.G., A.W.C., S.J.K., A.K.W. and J.A.J), Emergent Ventures Fast Grant (A.W.C.) and the Paul Ramsay Foundation (D.I.G., S.J.K., A.K.W and A.W.C). A.K.W., J.A.J, D.I.G., W.-H.T., S.J.K., and A.W.C are supported by NHMRC fellowships. W-H.T. is a Howard Hughes Medical Institute–Wellcome Trust International Research Scholar (208693/Z/17/Z). N.A.G. is supported by an ARC DECRA fellowship.

## Notes

### Competing Interest Statement

The authors have declared no competing interest.

### Author Declarations

The study protocols were approved by the University of Melbourne Human Research Ethics Committee (#2056689) and all associated procedures were carried out in accordance with the approved guidelines.

