## Supplementary material for "Heterologous SARS-CoV-2 IgA neutralising antibody responses in convalescent plasma": Supp materials

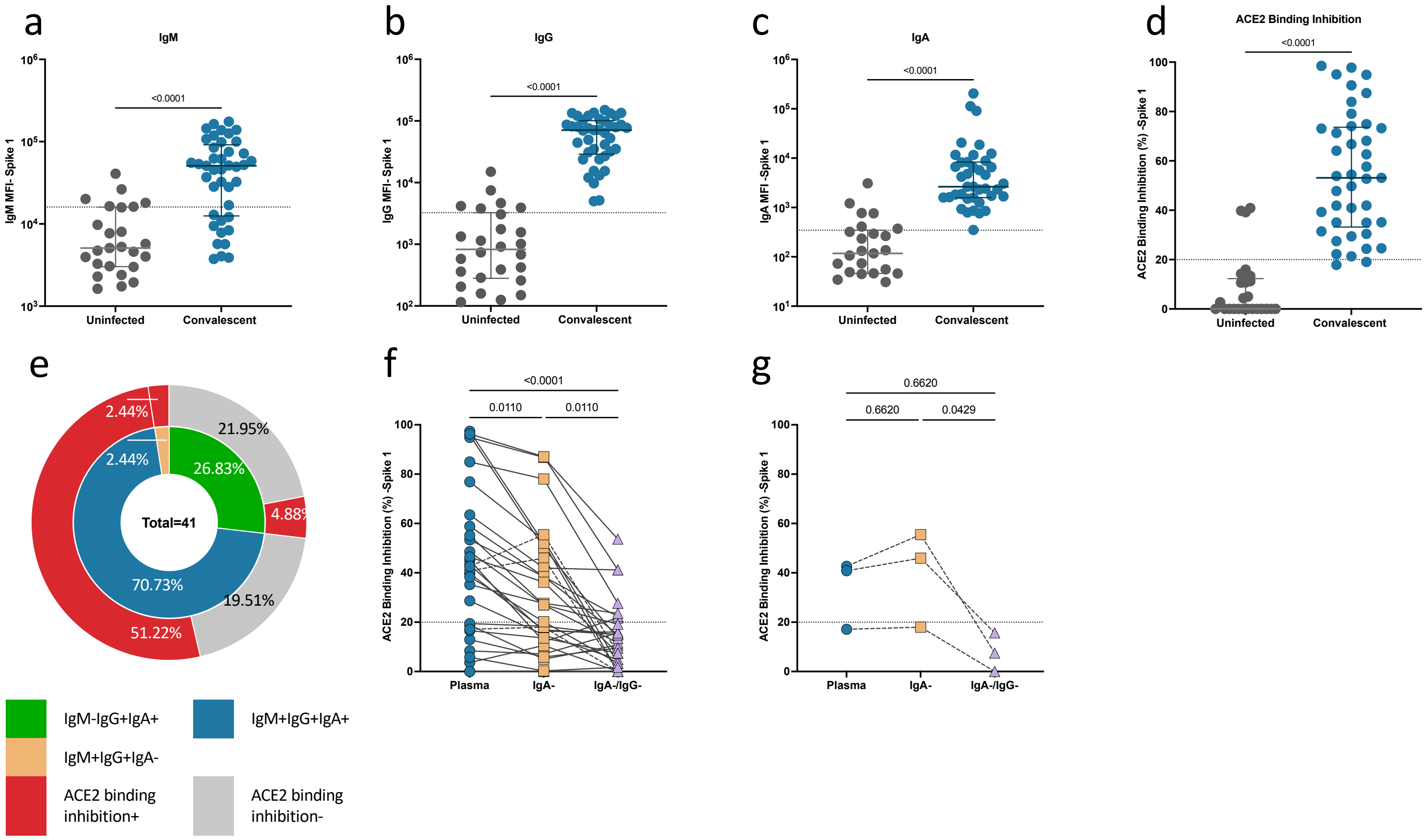


**Supplementary Figure 1. Anti-spike 1 plasma, IgA depleted plasma, IgA and IgG depleted plasma and purified IgG and IgA antibody isotype and ACE2 binding inhibition data**

Convalescent (n-41) (blue) and uninfected control (n=26) (grey) plasma (final dilution 1:100) was assessed for IgM **(a)**, IgG **(b)** and IgA **(c)** antibody binding to SARS-CoV-2 spike-1 (S1) via multiplex. A positive threshold (grey dotted line) was defined as the 75$^{th}$ percentile of antibody binding (MFI) for uninfected control plasma to respective antigens. **(d)** S1-ACE2 binding inhibition (%) of convalescent (blue) and uninfected control (grey) plasma (diluted 1:100). Statistical analyses between two groups were performed with the Mann-Whitney test. A positive threshold (grey dotted line) was defined as >20% ACE2 binding inhibition. **(e)** Pie chart outlining the percentage of subjects seropositive for anti-S1 antibody isotypes (IgM-IgG+IgA+ (green), IgM+IgG+IgA+ (blue), IgM+IgG+IgA- (yellow)) in the inner ring and the percentage of each seropositive subset with ACE2 binding inhibition in the outer red ring. SARS-CoV-2 S1 ACE2 binding inhibition (%) of **(f)** convalescent plasma (diluted 1:200; blue) and matched dilutions of IgA depleted (IgA-; yellow) and IgA plus IgG depleted (IgA-/IgG-; purple) (n=30) plasma fractions. Comparisons between plasma and depleted fractions was conducted using Friedman with Dunn's multiple comparison test. (sup. Fig. 3 describes inclusion/exclusion criteria for successful antibody depletion). **(g)** Samples where IgA depletion resulted in an increase in RBDwt ACE2 binding inhibition (n=3).

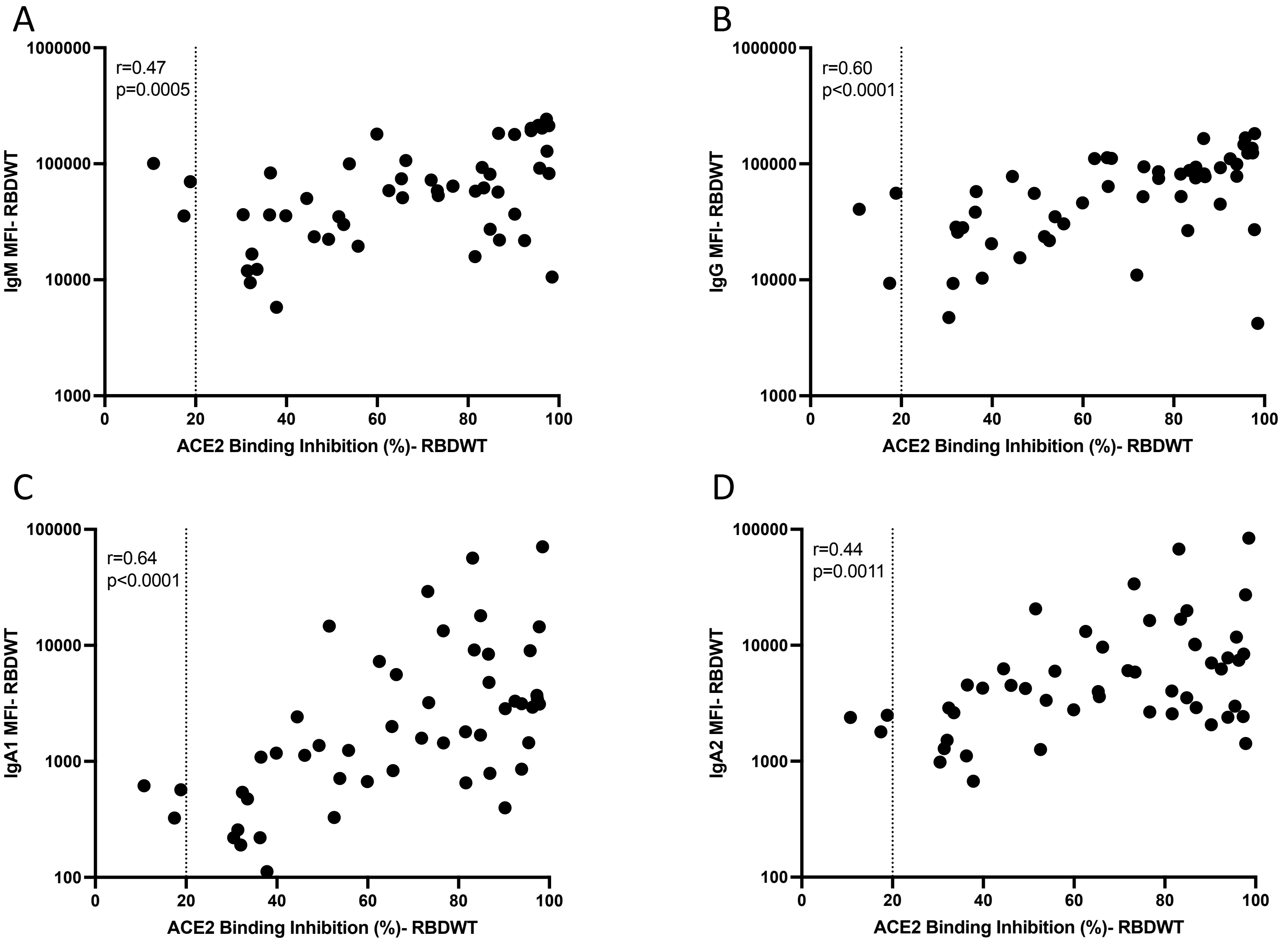


**Supplementary Figure 2. Individual correlations of convalescent RBDWT plasma IgM (a), IgG (b), IgA1 (c) and IgA2 (d) antibody binding with RBDWT-ACE2 binding inhibition**

### Supplementary Figure 3. Workflow of sample processing and confirmation of IgG and IgA depletion

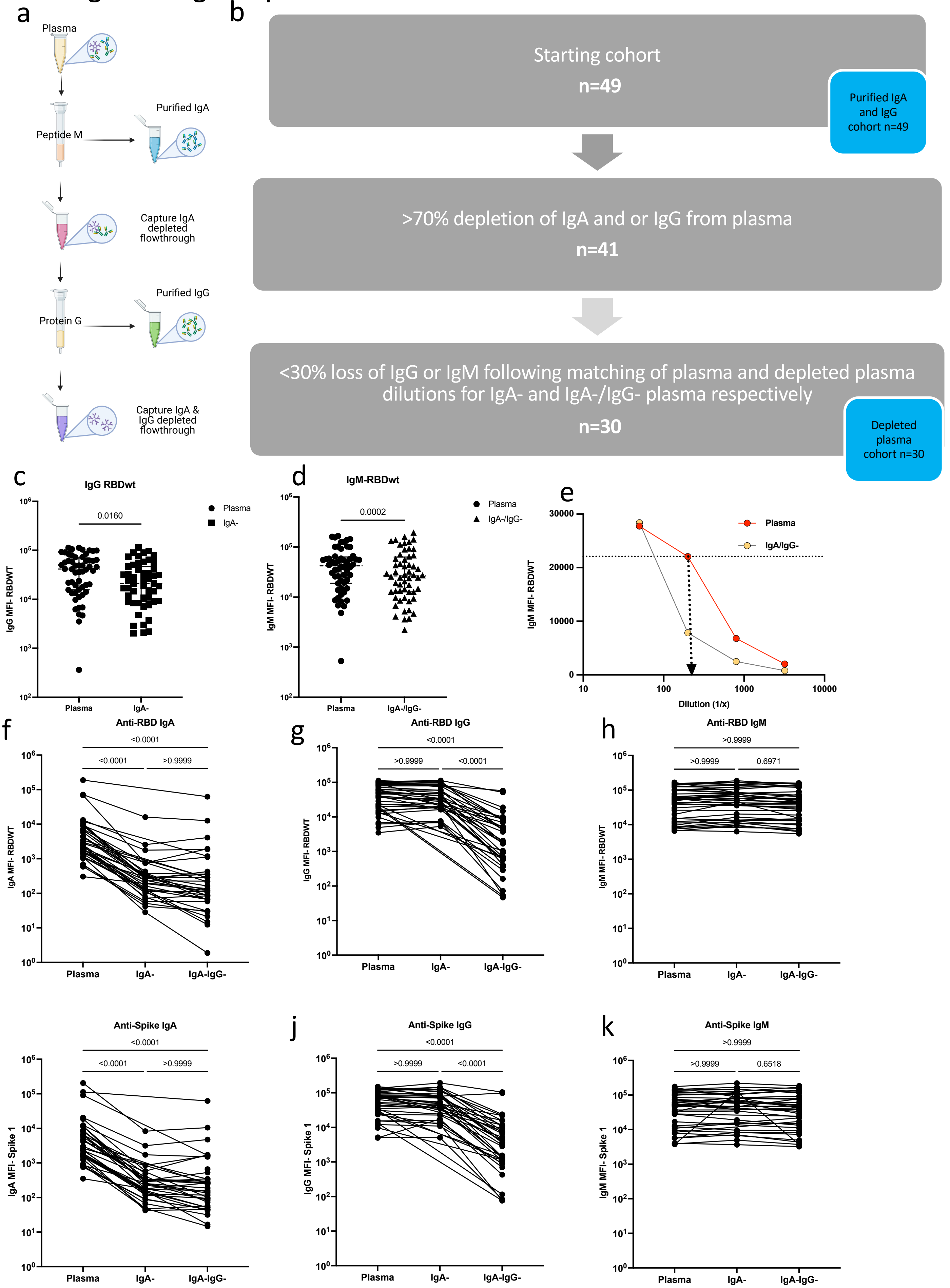

**Supplementary Figure 3. Workflow of sample processing and confirmation of IgG and IgA depletion**
**(a)** Depletion and purification of IgA and IgG from convalescent plasma and **(b)** sample exclusion process. **(c-d)** Significant loss of anti-RBDWT IgG and IgM from IgA- and IgA-/IgG- plasma respectively (1:100 dilution). For fair comparison, depleted plasma dilutions were matched to complete plasma (1:100 dilution). Using anti RBDWT IgG or IgM binding MFI of IgA- and IgA-/IgG- plasma respectively **(e)**, a plasma dilution was selected to match the antibody MFI for complete plasma (yellow) at 1:100 dilution. Matching is indicated by the black dotted arrow. Anti-RBDWT IgA **(f)**, IgG **(g)** and IgM **(h)** MFI for plasma, IgA- and IgA and IgA-/IgG- for matched dilutions. Anti-Spike 1 IgA **(i)**, IgG **(j)** and IgM **(k)** MFI for plasma, IgA- and IgA and IgA-/IgG- for matched dilutions.

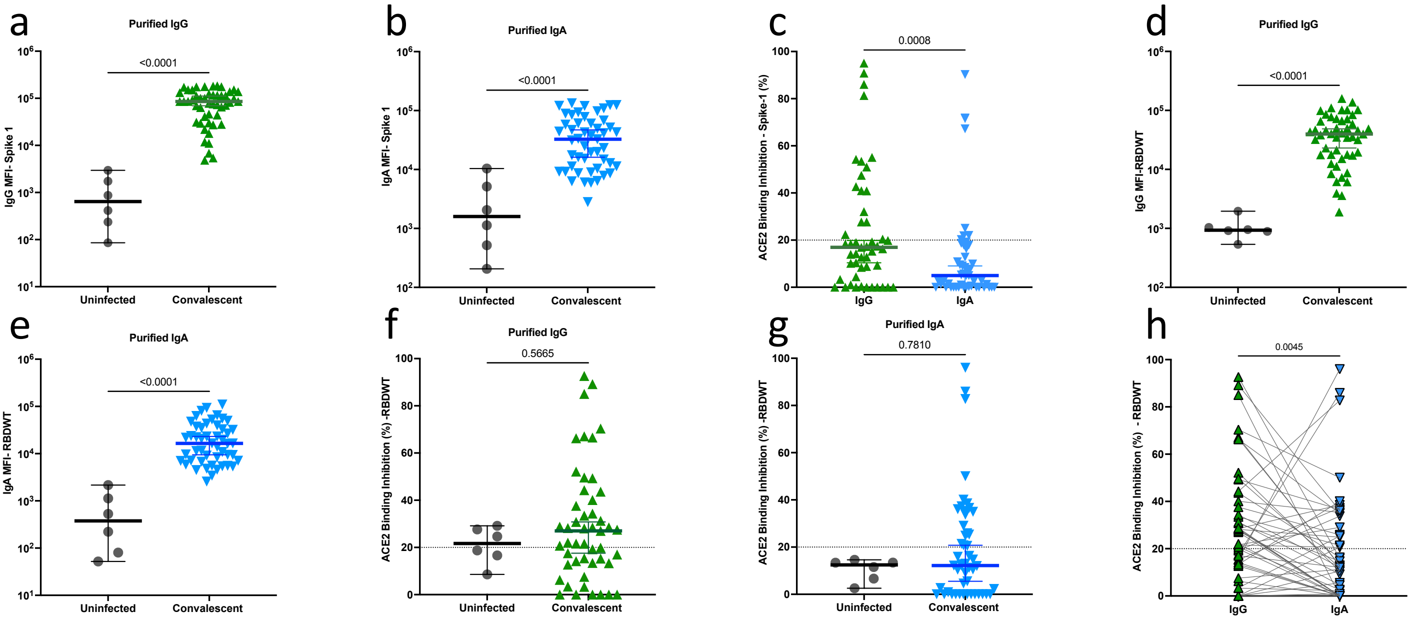


**Supplementary Figure 4. Purified convalescent IgG and IgA spike 1 and RBDWT supplementary data**
Purified IgA **(a)** (blue) and IgG **(b)** (green) anti-S1 antibody binding (MFI) for uninfected (n=6, grey) and convalescent (n=49) cohorts at 100ug/ml total antibody. **(c)** Purified IgA (blue) and IgG (green) S1-ACE2 binding inhibition (%) at 100ug/ml total antibody (n=49). Purified IgA **(d)** (blue) and IgG **(e)** (green) anti-RBDWT antibody binding (MFI) for uninfected (n=6, grey) and convalescent (n=49) subjects at 100ug/ml total antibody. Purified IgA **(f)** (blue) and IgG **(g)** (green) anti-RBDWT RBDWT-ACE2 binding inhibition for uninfected (n=6, grey) and convalescent (n=49) cohorts at 100ug/ml total antibody. **(h)** Purified IgA (blue) and IgG (green) RBDWT-ACE2 binding inhibition (%) at 100ug/ml total antibody (n=49). Lines connect IgG and IgA for a single individual.
A positive threshold of 20% ACE2 binding inhibition was set (grey dotted line).

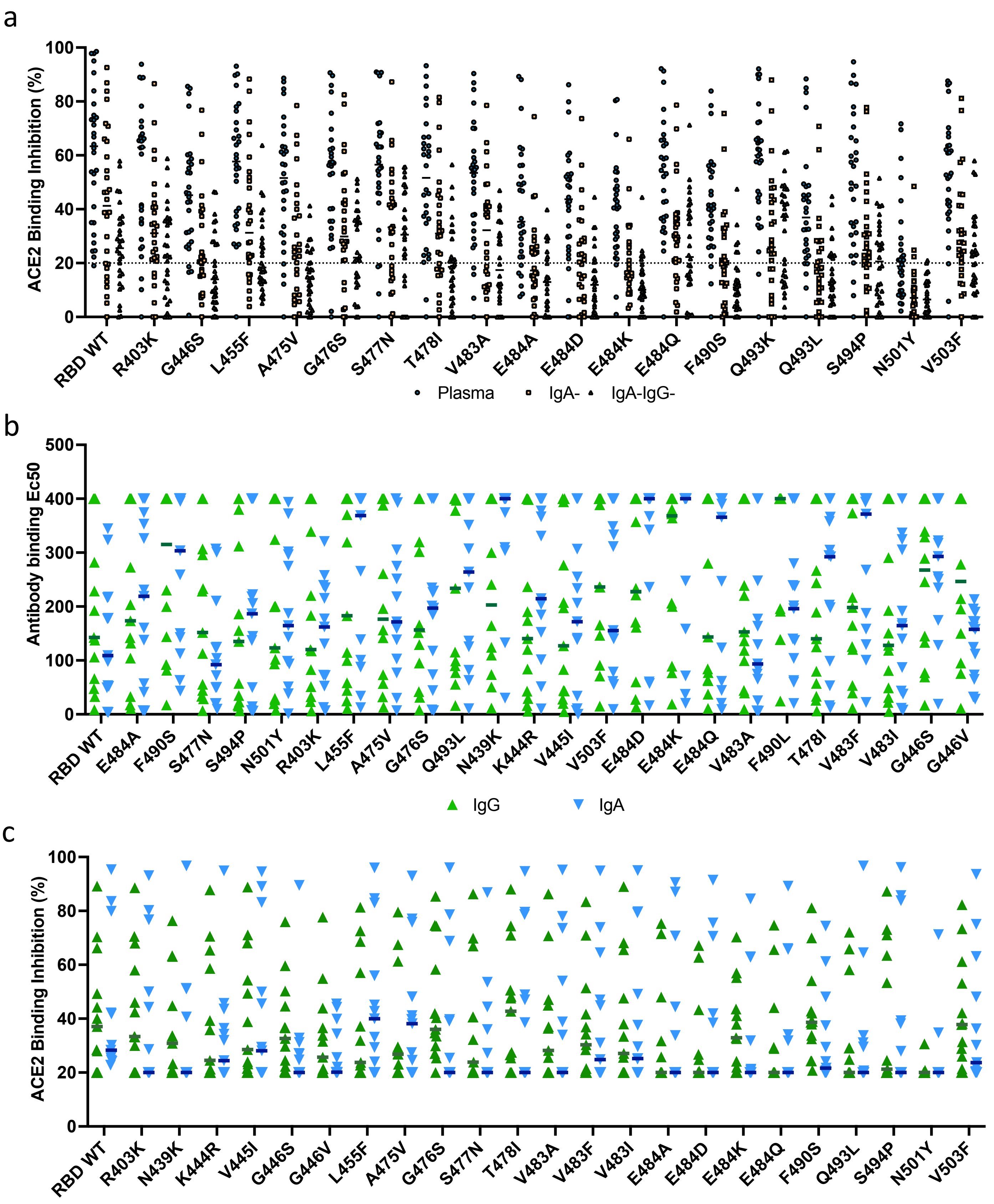


**Supplementary Figure 5. Raw ACE2 binding inhibition and antibody binding data for depleted plasma and purified antibodies to single RBD mutations.**
**(a)** ACE2 binding inhibition (%) to RBDWT and 18 other RBD single mutants for plasma (blue), IgA depleted (IgA-, yellow) plasma and IgA and IgG depleted (IgA-IgG-, purple triangle) plasma. Raw purified IgG (green) and IgA (blue) Ec50's **(b)** and ACE2 binding inhibition (%) **(c)** for RBDWT and 23 other RBD single mutants.
Lines depict the median response for each sample type to each RBD.

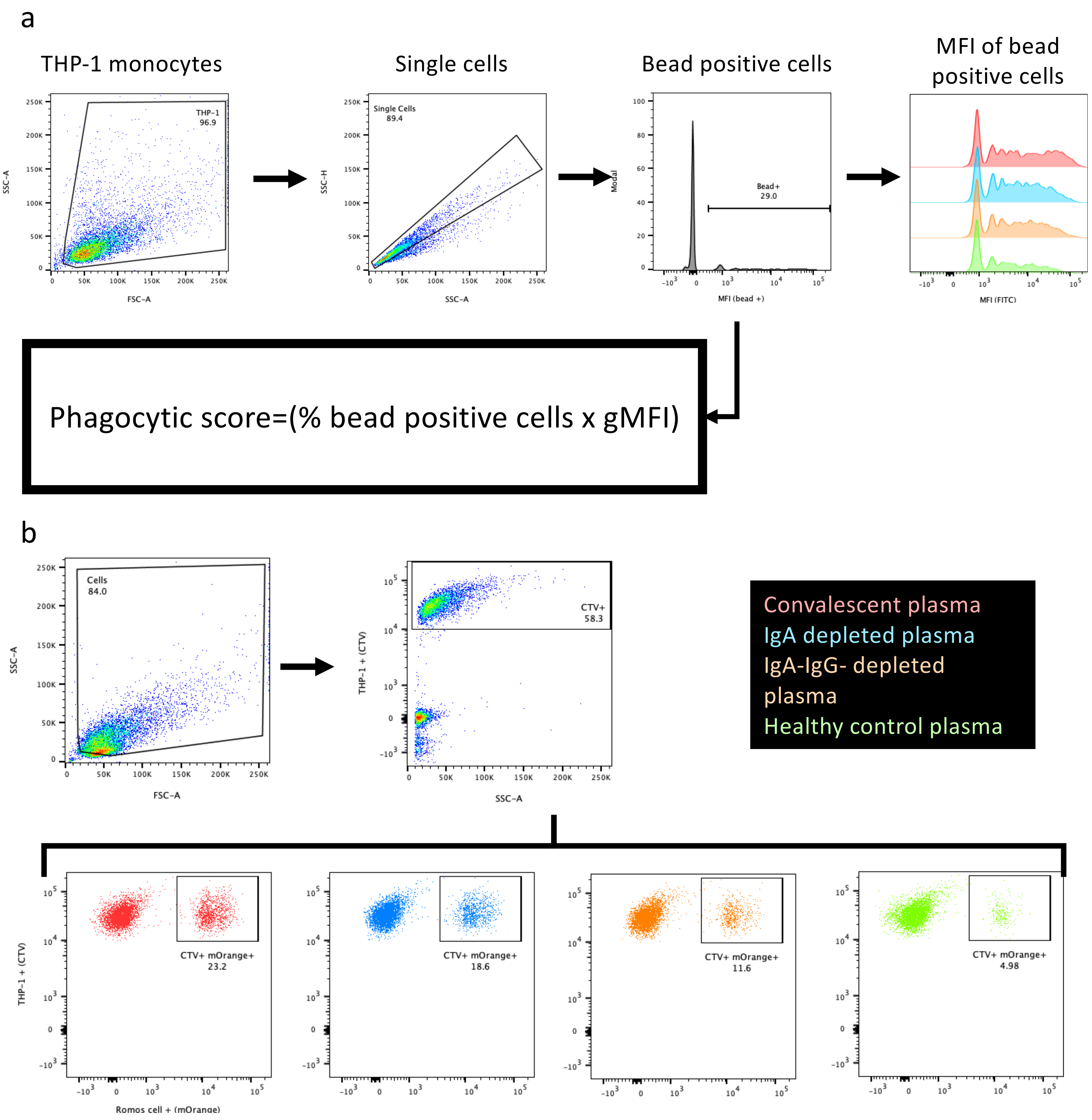


**Supplementary Figure 6. Gating strategy of ADCP bead-based assay and THP-1 Ramos S-orange association assay.**
**(a)** Antibody dependent bead-based phagocytosis assay gating strategy. First, THP-1 monocytes were gated, followed by single cells and bead positive cells. Finally, a phagocytic score was calculated. **(b)** THP-1 and Ramos S-orange cell association assay gating strategy. Cells were gated, followed by cell trace violet (CTV) stained THP-1 monocytes. Finally, CTV+ (THP-1) mOrange+ (Ramos S-orange) cells were gated for and the percentage of double positives was recorded as a measure of anti-spike antibody mediated cell association.
Example convalescent plasma (red), IgA- plasma (blue), IgA-/IgG- plasma (orange) and healthy control plasma (green) are shown.

**Supplementary table S1: Subject demographics**

| | | Healthy controls | | COVID-19 Convalescent | |
|---|---|---|---|---|---|
| | | Multiplex (n=26) | Functional (n=12)* | Multiplex (n=41) | Functional (n=39)* |
| Sample collection data (month) | | April-May 2020 | April-May 2020 | April-May 2020 | April-May 2020 |
| Date of PCR test (month) | | | | March 2020 | March 2020 |
| Age, Median (IQR) | | 54 (24-60) | 55 (24-63) | 55 (49-61) | 57 (52-69) |
| Gender, female (%) | | 12 (48) | 5 (45.45) | 11 (26.83) | 11 (28.21) |
| PCR and/or serology | | Negative** | Negative** | Positive | Positive |
| Disease Severity (self-report) | Mild (%) | - | - | 27 (65.85) | 26 (68.42) |
| | Moderate (%) | - | - | 7 (17.07) | 6 (15.79) |
| | Sever (%) | - | - | 2 (4.88) | 1 (2.63) |
| | No details (%) | - | - | 5 (12.20) | 5 (13.16) |
| Days post positive COVID test, median (IQR) | | - | - | 36 (30-44) | 36 (30-38) |
| Days post symptom onset, median (IQR) | | - | - | 41 (36-47) | 40 (38-42) |

- Not applicable

*Individuals were randomly selected from original cohort

**Confirmed negative using serology

**Supplementary Table 2. Matched depleted plasma dilution to completes plasma at 1:100**

| | Subject ID | **Dilution IgA- (1/n)** | **Dilution IgA-/IgG- (1/n)** |
|---|---|---|---|
| 1 | CP07 | 100 | 100 |
| 2 | CP11 | 50 | 50 |
| 3 | CP15 | 50 | 50 |
| 4 | CP19 | 100 | 100 |
| 5 | CP23 | 100 | 100 |
| 6 | CP25 | 100 | 100 |
| 7 | CP26 | 100 | 100 |
| *8* | CP27 | 50 | 50 |
| 9 | CP29 | 100 | 100 |
| 10 | CP30 | 100 | 50 |
| 11 | CP31 | 50 | 50 |
| 12 | CP32 | 50 | 100 |
| 13 | CP35 | 100 | 50 |
| 14 | CP39 | 100 | 100 |
| 15 | CP43 | 100 | 100 |
| 16 | CP44 | 100 | 100 |
| 17 | CP45 | 100 | 100 |
| 18 | CP46 | 100 | 100 |
| 19 | CP51 | 100 | 100 |
| 20 | CP52 | 50 | 50 |
| 21 | CP53 | 50 | 50 |
| 22 | CP54 | 50 | 100 |
| 23 | CP57 | 100 | 100 |
| 24 | CP58 | 50 | 100 |
| 25 | CP60 | 100 | 100 |
| *26* | CP61 | 100 | 100 |
| *27* | CP63 | 100 | 100 |
| 28 | CP66 | 50 | 50 |
| 29 | CP68 | 50 | 100 |
| 30 | CP70 | 50 | 50 |
